# Household Size and Age as Primary Drivers of COVID-19 Infection Among Priority Populations in Australia

**DOI:** 10.64898/2026.03.23.26349117

**Authors:** Shanti Narayanasamy, Aimée Altermatt, Wai Chung Tse, Lisa Gibbs, Anna L. Wilkinson, Katherine Heath, Mark Stoové, Nick Scott, Katherine B. Gibney, Margaret Hellard, Alisa Pedrana

## Abstract

**Background:** The COVID-19 pandemic exacerbated health disparities globally, with certain populations experiencing disproportionate disease burdens. In Australia, COVID-19 deaths occurred disproportionately among first-generation migrants. This study examined risk factors for COVID-19 infection in a Victorian cohort recruited from priority populations, including healthcare workers, people with chronic health conditions, and culturally and linguistically diverse (CALD) communities.

**Methods:** We conducted a cross-sectional analysis of participants from the Optimise longitudinal cohort study (September 2020–December 2023). The primary outcome was the self-reported count of confirmed COVID-19 infections (PCR or rapid antigen test positive) from December 2019 to December 2023. We used Poisson regression to examine associations between baseline sociodemographic characteristics and infection count, calculating unadjusted and adjusted incidence rate ratios (IRRs) with 95% confidence intervals (CIs).

**Results:** Of 433 participants (median age 51 years, 75% female), 25% reported no infections, 48% reported one infection, and 27% reported two or more infections. In univariate analysis, CALD status (IRR=1.24,95%CI:1.02–1.50) and larger household size (2-5 people, IRR=1.71,95%CI:1.14-2.50) were associated with higher infection rates, while chronic health conditions (IRR=0.73, 95%CI:0.61–0.88) and older age (IRR=0.54, 95%CI:0.43–0.67) were associated with lower infection rates. In adjusted analysis, younger age (18-34 years vs ≥55 years: aIRR=0.63,95%CI:0.48–0.82) and medium household size (living alone vs 2-5 person household: aIRR=1.42, 95%CI:1.11–1.83) remained significant predictors. CALD status and socioeconomic status showed no independent association with infection risk after adjustment for household size and age.

**Conclusion:** COVID-19 infection risk in this Victorian cohort was driven by younger age and larger household size rather than CALD status or socioeconomic status, suggesting that housing density and age, rather than cultural or socioeconomic characteristics, determined infection patterns. Future pandemic preparedness should prioritise policies enabling safe quarantine and isolation for individuals in larger households and workplace protections and economic security for younger essential workers.

## BACKGROUND

The COVID-19 pandemic exacerbated longstanding health and social disparities globally. In Victoria, Australia, several groups have been identified at higher risk of COVID-19 infection and mortality: frontline workers, people experiencing unemployment or housing stress, individuals living in higher density housing, those speaking languages other than English at home and overseas-born residents.(1, 2) While Australia’s COVID-19 mortality was substantially lower than similarly resourced nations such as the United States and United Kingdom, most Australian COVID-19 deaths occurred disproportionately among first-generation migrants; there were 2.3 COVID-19 deaths per 100,000 people born in Australia, compared with 6.8 for those born overseas.(3–5) During the pandemic infection numbers varied across different waves, with most cases occurring during the Omicron wave (late 2021 to early 2022).(6)

Understanding the determinants of single and repeated infections is important to identify which communities may be at higher risk for infection, and why. While single infections were common across the population, multiple infections may indicate distinct risk profiles related to occupation, household transmission or behavioural risks. Sociodemographic characteristics such as age, household composition, socioeconomic status (SES), and cultural background have been identified as key determinants of COVID-19 risk, yet their relative contributions to infection patterns, particularly repeated infections, remain incompletely understood.

This study aims to characterise the risk factors for COVID-19 infection among the Optimise study cohort. The Optimise study recruited from priority and at-risk populations: healthcare workers, aged care workers (and other at-risk occupations), people with pre-existing chronic health conditions, people speaking a language other than English at home, and people from culturally and linguistically diverse communities (CALD). By characterising infection patterns across this intentionally diverse cohort, we aim to identify which population subgroups experienced disproportionate infection burdens and whether these disparities persisted after accounting for sociodemographic and occupational factors. Such insights are critical for informing future targeted public health interventions and understanding how pandemic impacts were distributed across Victorian communities, particularly among groups that faced elevated risks due to structural inequalities or occupational exposures.

The primary aim of this study is to describe the participants with COVID-19 infection in the Optimise cohort. The secondary aim is to describe the risk factors for one or multiple (>1) COVID-19 infections during the study period.

## METHODS

### Study design

The Optimise Study (Optimise) was a mixed-methods longitudinal cohort and social networks study (September 2020–December 2023) that aimed to assess the impact of the COVID-19 pandemic and public health measures on the community in Victoria, Australia.(7) Participants aged ≥18 years were recruited between 27 September 2020 and 18 December 2021; informed verbal consent was obtained. Optimise was created to obtain insights into priority populations deemed at-risk of: (i) acquiring COVID-19; (ii) experiencing severe COVID-19 illness; or (iii) suffering negative impacts from pandemic-related restrictions.(7) Participants were recruited via both paid and organic social media campaigns, as well as flyers distributed through community and industry organisations, community-based groups and social and professional networks. Additionally, the study employed bilingual research personnel to connect with Mandarin-, Arabic-, and Dinka-speaking individuals, and study promotional materials were translated into Mandarin and Arabic, with surveys available for completion in English, Mandarin and Arabic. A detailed description of the study is available elsewhere.(7) We used a cross-sectional study design to describe the participants with COVID-19 infection in the Optimise cohort and understand the risk factors for multiple COVID-19 infections.

### Setting

Victoria was under a State of Emergency from March 20, 2020 to December 21, 2021. During that time there were five lockdown periods during which three broad categories of public health measures were enacted to control the transmission of SARS-CoV-2 – public health measures that were not enforced, others that were required and COVID-19 testing and isolation requirements for individuals with flu-like symptoms or COVID-19 contacts.(8)

During Victoria’s State of Emergency period, individuals were required to test for COVID-19 if they had cold or flu symptoms or were identified as a close contact of someone with COVID-19. When the pandemic declaration ended in Victoria (12 October 2022), this mandatory testing requirement was removed, and COVID-19 testing became a recommendation rather than a requirement for symptomatic individuals or close contacts.(9)

COVID-19 testing modality also changed in Victoria during the study period. SARS-CoV-2 polymerase chain reaction (PCR) tests were the only method of testing during the early pandemic period. Victoria made rapid antigen tests (RATs) widely available from late 2021, and integrated large-scale distribution and reporting into the public health system by early 2022.(10, 11) By March 2022, RATs dominated household testing: of households that took a COVID-19 test that month, 90% used a RAT.(12)

### Data sources

This study uses the following data sources from Optimise: 1) a baseline survey (comprehensive survey on enrolment that captured sociodemographic information, physical health status and caring responsibilities both before and during the COVID-19 pandemic); 2) follow-up surveys (monthly surveys completed every 28 days collecting similar information as the baseline survey, with past four weeks recall); and 3) follow-up diaries (collected up to four times during each month to capture data on COVID-19 testing, symptoms and mental health in the past seven days), 4) the 2023 follow up survey (conducted in December 2023) a standalone survey capturing sociodemographic data, health status and COVID-19 infection-related data.

### Participants

Individuals were eligible to participate in Optimise if they were aged ≥18 years, lived in Victoria, and had internet or phone access. All participants received monthly electronic gift vouchers as reimbursement for their participation.

### Inclusion criteria

To be included in this study participants had to have 1) completed the 2023 follow-up survey, 2) have completed the baseline survey with data for postcode and language spoken at home, and 3) not have missing data for key variables (baseline demographics, chronic disease status, household income and education status). Responses of ‘prefer not to say’ or ‘don’t know’ were classified as missing.

### Outcomes

The primary outcome was the count of self-reported confirmed COVID-19 infections between December 2019 and December 2023, collected via the 2023 follow-up survey. Participants were asked: “*How many times have you been infected with confirmed COVID-19 since the start of the pandemic (December 2019)?*”. A confirmed COVID-19 infection was defined as participants testing positive by PCR or RAT, with at least one month between successive positive tests. Responses reporting more than 20 infections were considered data entry errors, treated as missing data and excluded from analysis.

To address recall bias, we cross-validated self-reported infection counts against data collected prospectively during the study period using a mix of follow-up survey and diaries. (refer to Supplementary Methods for details).

### Covariates

We examined pre-pandemic demographic, socioeconomic, and health-related characteristics collected at baseline, including: age; sex; language spoken at home (English versus other); country of birth (Australia versus overseas); parental country of birth (Australia versus overseas); residential postcode; employment status; work environment (home-based versus other); household income (prior to the COVID-19 pandemic); household size (number of adults and children); presence of chronic health conditions (including immunocompromised status); residence type (owner-occupied versus other); use of childcare services; and unpaid carer status.

Participants were classified in the CALD population if a) their country of birth was not Australia, the United Kingdom or Ireland, or b) their language spoken at home was not English. The CALD definition used for this study reflects the population in Australia with the highest mortality from COVID-19.(5)

Socio-economic groups were defined based on the SEIFA (Socio-Economic Indexes for Areas) Index of Relative Socio-Economic Disadvantage (IRSD) based on postcode at baseline.(^13^)^,6^ Participants were classified in the low-SES group if they resided in an area classified in IRSD deciles 1–6, and high-SES group by IRSD deciles 7–10.

### Statistical Methods

We used descriptive analyses to compare socio-demographics at baseline by number of COVID-19 infections, categorised into zero, one infection or two or more infections.

Multivariate Poisson regression was used to examine the relationship between baseline characteristics and the outcome of count of COVID-19 infections, without an offset for exposure time. The outcome data were approximately equi-dispersed, making a Poisson regression appropriate. We estimated unadjusted and adjusted incidence rate ratios (IRRs) with 95% confidence intervals and global p-values. The multivariable model included all variables that were potential protective factors or risk factors for COVID-19 infection.

All statistical analyses were performed using R version 4.4.1 with statistical significance set at p<0.05.

### Sensitivity Analysis

We treated the work environment as a stable variable in the main analysis, using baseline survey responses. However, as we expected this variable could fluctuate over time due to changing public health restrictions, we conducted a sensitivity analysis that reclassified participants based on their work environment patterns across the data collected throughout the study period. Participants were classified into three groups based on work environment: 1) those who always worked from home, 2) those who always attended the workplace, and 3) those who shifted between working from home and the workplace. We expected participants who always attended the workplace to have a higher incidence of COVID-19.

### Patient and Public Involvement

COVID-19 patients and members of the public were involved in this research, a detailed description is noted elsewhere.(7)

### Ethics

Ethics approval for Optimise was provided by the Alfred Human Research Ethics Committee, Approval Number 333/20.

## RESULTS

Optimise recruited 779 participants in total between September 2020 and September 2021: 558 (72%) completed the 2023 follow-up survey. Of this group, 31 (5.6%) were removed for missing outcome data, 82 (14.7%) were removed for missing baseline data on age, postcode, gender, chronic health condition, education, household income or carer status and two (0.4%) were removed because their reported number of infections were deemed to be outliers (≥20) (**Figure 1**). We explored the option to impute data from follow-up surveys, however because missingness involved key baseline covariates and could not be reliably predicted from later measurements. The final analysis dataset contained 443 participants.

**Figure 1:**
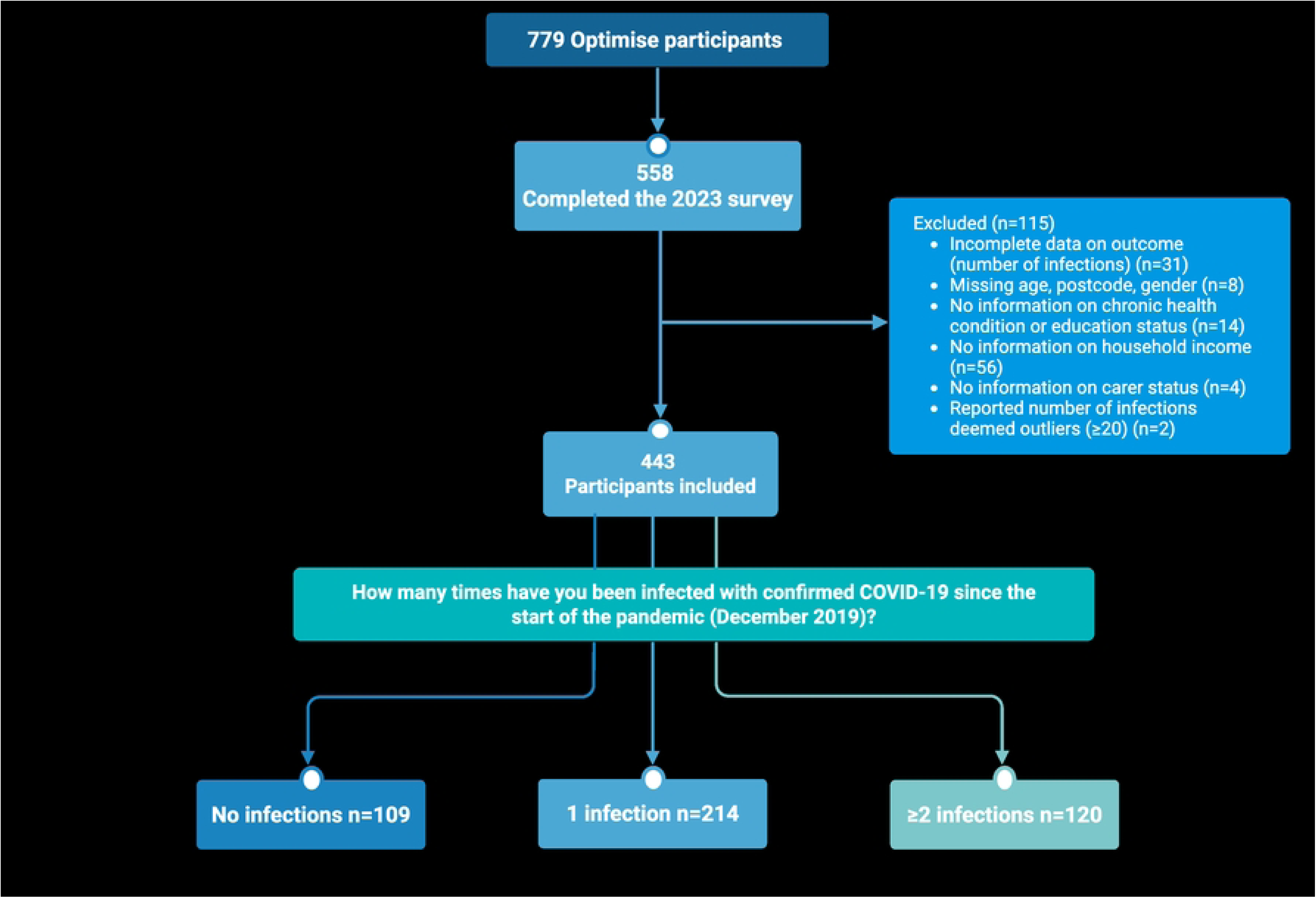
Flowchart of participants included in this study.

### Descriptive data

Participants (n=433) were aged 23–89 years (median age 51, IQR 36–65). Most were women (75%), spoke English as the main language at home (86%), were born in Australia (67%) and were residing in a postcode with classified as high SES at baseline (55%) (**Table 1**).

**Table 1:**
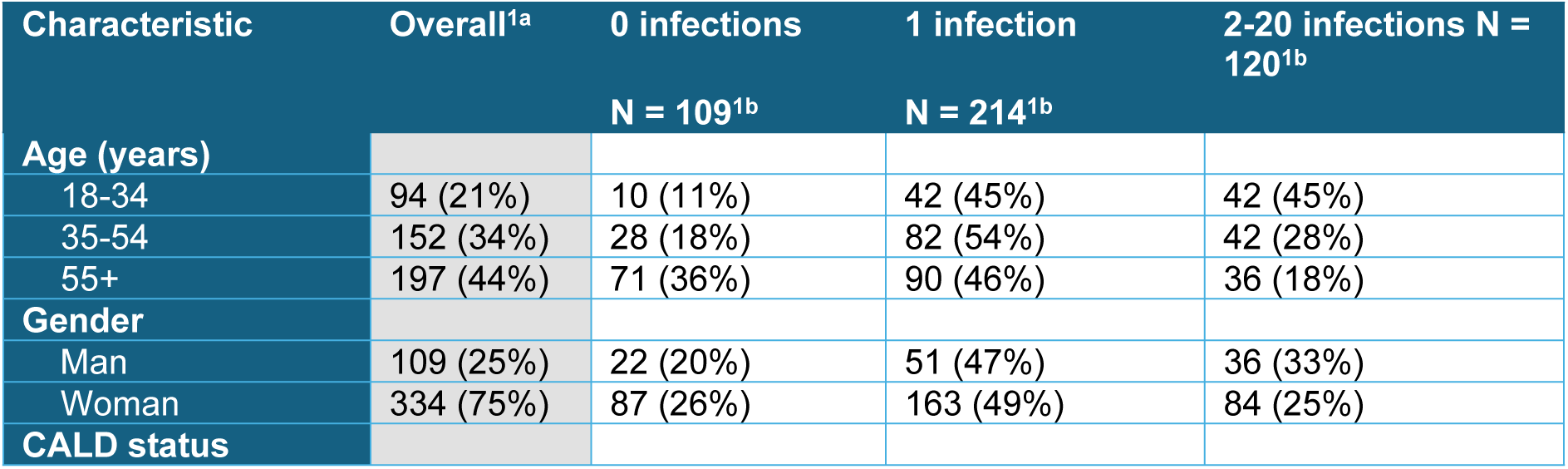
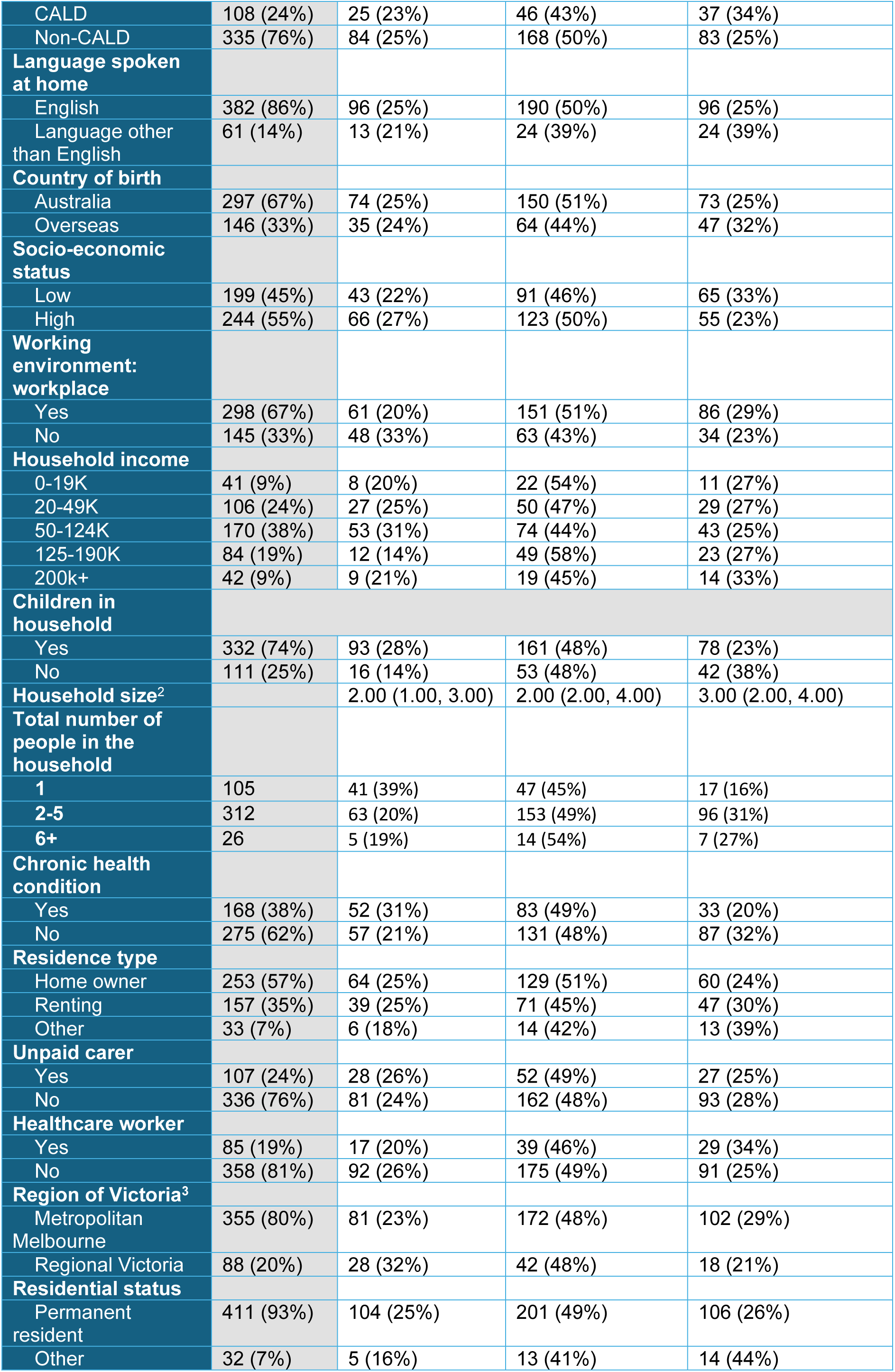
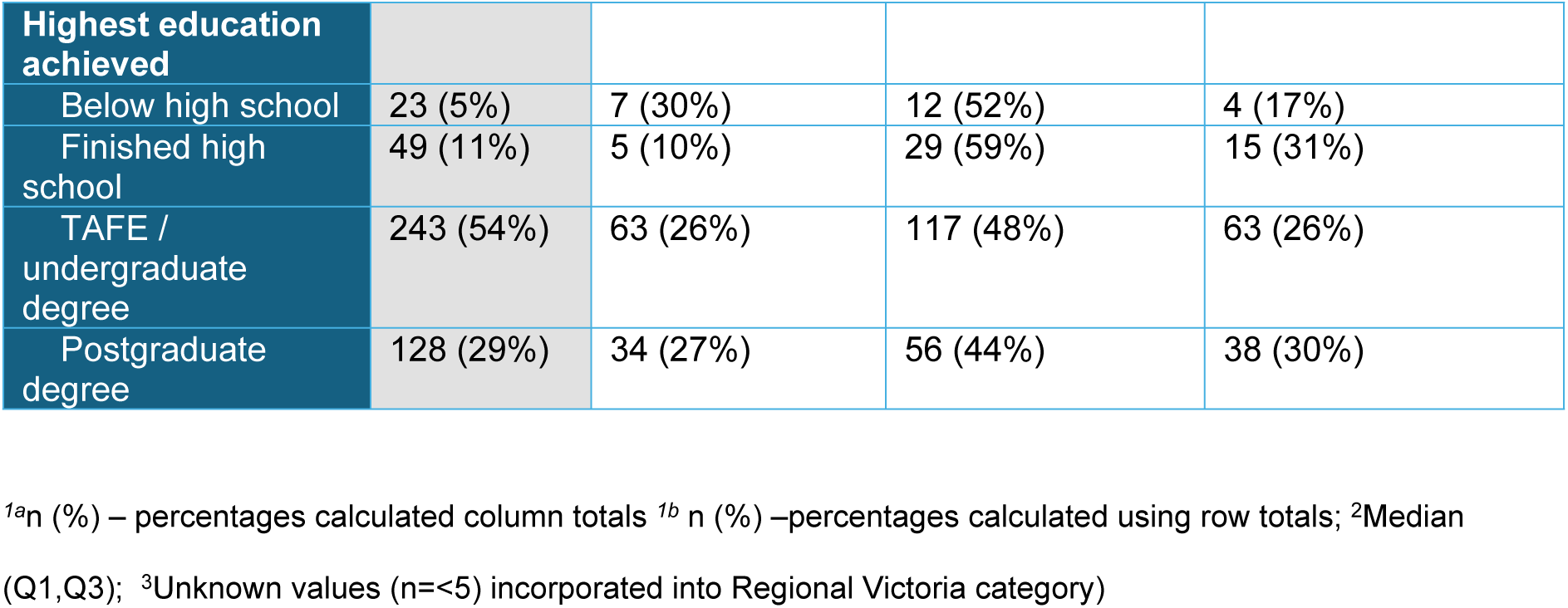
Sociodemographic characteristics of Optimise study participants at baseline survey (N=443).

One hundred and nine (25%) participants reported no COVID-19 infection, 214 (48%) reported one infection and 120 (27%) reported two or more infections over the study period (see Supplementary Results for further details). In the CALD group, 23% had zero infection, 43% reported one infection and 34% reported two or greater infections, compared with the non-CALD group who reported 25% (zero infections), 50% (one infection) and 25% two or more infections.

### Main Results - Univariate Analysis

The outcome for count of COVID-19 infections was approximately equi-dispersed (mean 1.15 infections, variance 0.98). In the unadjusted Poisson regression analysis, several baseline characteristics were significantly associated with COVID-19 infection count (**Table 2**). CALD participants had a 24% higher rate of infection compared to non-CALD participants (IRR=1.24,95% CI:1.02–1.50,p=0.032). Age showed a strong inverse association with infection rate (p<0.001), with participants aged 35–54 years having a 22% lower infection rate (IRR=0.78, 95% CI:0.63–0.96) and those aged 55+ years having a 46% lower infection rate (IRR=0.54, 95% CI:0.43–0.67) compared to the 18–34 age group.

**Table 2:**
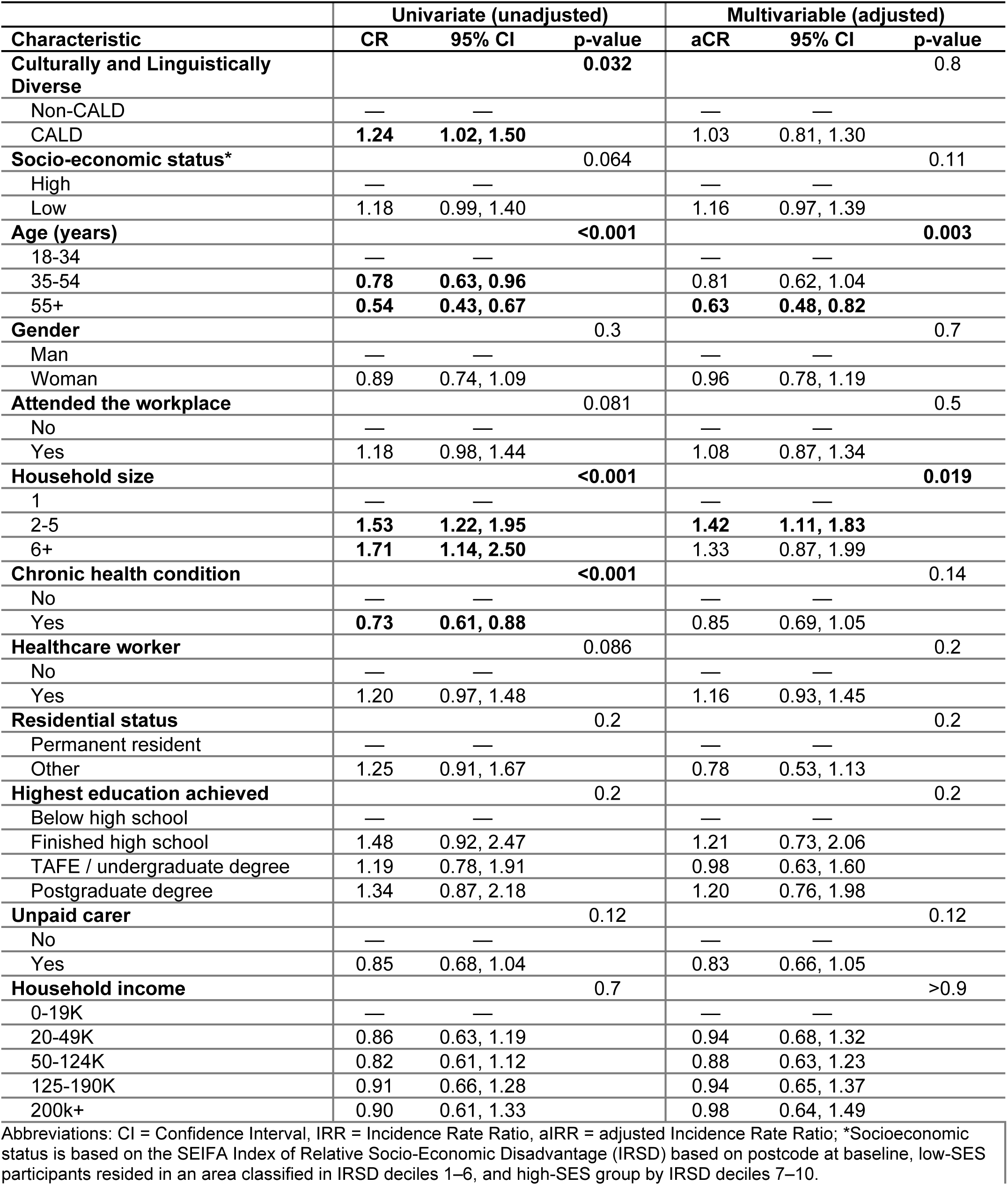
Unadjusted and Adjusted Incidence Rate Ratios (IRRs) for any COVID-19 infection, using baseline characteristics (N=443), 2019–2023.

| Characteristic | Univariate (unadjusted) |  |  | Multivariable (adjusted) |  |  |
| --- | --- | --- | --- | --- | --- | --- |
|  | CR | 95% CI | p-value | aCR | 95% CI | p-value |
| **Culturally and Linguistically Diverse** |  |  | **0.032** |  |  | **0.8** |
| Non-CALD | — | — |  | — | — |  |
| CALD | **1.24** | **1.02, 1.50** |  | 1.03 | 0.81, 1.30 |  |
| **Socio-economic status\*** |  |  | 0.064 |  |  | 0.11 |
| High | — | — |  | — | — |  |
| Low | 1.18 | 0.99, 1.40 |  | 1.16 | 0.97, 1.39 |  |
| **Age (years)** |  |  | **<0.001** |  |  | **0.003** |
| 18-34 | — | — |  | — | — |  |
| 35-54 | **0.78** | **0.63, 0.96** |  | 0.81 | 0.62, 1.04 |  |
| 55+ | **0.54** | **0.43, 0.67** |  | **0.63** | **0.48, 0.82** |  |
| **Gender** |  |  | 0.3 |  |  | 0.7 |
| Man | — | — |  | — | — |  |
| Woman | 0.89 | 0.74, 1.09 |  | 0.96 | 0.78, 1.19 |  |
| **Attended the workplace** |  |  | 0.081 |  |  | 0.5 |
| No | — | — |  | — | — |  |
| Yes | 1.18 | 0.98, 1.44 |  | 1.08 | 0.87, 1.34 |  |
| **Household size** |  |  | **<0.001** |  |  | **0.019** |
| 1 | — | — |  | — | — |  |
| 2-5 | **1.53** | **1.22, 1.95** |  | **1.42** | **1.11, 1.83** |  |
| 6+ | **1.71** | **1.14, 2.50** |  | 1.33 | 0.87, 1.99 |  |
| **Chronic health condition** |  |  | **<0.001** |  |  | 0.14 |
| No | — | — |  | — | — |  |
| Yes | **0.73** | **0.61, 0.88** |  | 0.85 | 0.69, 1.05 |  |
| **Healthcare worker** |  |  | 0.086 |  |  | 0.2 |
| No | — | — |  | — | — |  |
| Yes | 1.20 | 0.97, 1.48 |  | 1.16 | 0.93, 1.45 |  |
| **Residential status** |  |  | 0.2 |  |  | 0.2 |
| Permanent resident | — | — |  | — | — |  |
| Other | 1.25 | 0.91, 1.67 |  | 0.78 | 0.53, 1.13 |  |
| **Highest education achieved** |  |  | 0.2 |  |  | 0.2 |
| Below high school | — | — |  | — | — |  |
| Finished high school | 1.48 | 0.92, 2.47 |  | 1.21 | 0.73, 2.06 |  |
| TAFE / undergraduate degree | 1.19 | 0.78, 1.91 |  | 0.98 | 0.63, 1.60 |  |
| Postgraduate degree | 1.34 | 0.87, 2.18 |  | 1.20 | 0.76, 1.98 |  |
| **Unpaid carer** |  |  | 0.12 |  |  | 0.12 |
| No | — | — |  | — | — |  |
| Yes | 0.85 | 0.68, 1.04 |  | 0.83 | 0.66, 1.05 |  |
| **Household income** |  |  | 0.7 |  |  | >0.9 |
| 0-19K | — | — |  | — | — |  |
| 20-49K | 0.86 | 0.63, 1.19 |  | 0.94 | 0.68, 1.32 |  |
| 50-124K | 0.82 | 0.61, 1.12 |  | 0.88 | 0.63, 1.23 |  |
| 125-190K | 0.91 | 0.66, 1.28 |  | 0.94 | 0.65, 1.37 |  |
| 200k+ | 0.90 | 0.61, 1.33 |  | 0.98 | 0.64, 1.49 |  |

Abbreviations: CI = Confidence Interval, IRR = Incidence Rate Ratio, aIRR = adjusted Incidence Rate Ratio; \*Socioeconomic status is based on the SEIFA Index of Relative Socio-Economic Disadvantage (IRSD) based on postcode at baseline, low-SES participants resided in an area classified in IRSD deciles 1–6, and high-SES group by IRSD deciles 7–10.

Household size demonstrated a significant positive association with infection rate (p<0.001). Participants living in households of 2–5 people had a 53% higher rate of infections (IRR=1.53,95% CI:1.22–1.95), while those in households of 6 or more people had a 71% higher rate (IRR=1.71,95% CI:1.14–2.50) compared to those living alone. Conversely, participants with chronic health conditions had a 27% lower infection rate than those without (IRR=0.73,95% CI:0.61–0.88,p< 0.001).

### Main Results – Multivariate Analysis

In the multivariable model age remained a significant predictor of infection (p=0.003) with participants aged over 55 years having a 37% lower infection rate compared to those aged 18–34 years (aIRR=0.63,95% CI: 0.48–0.82), while the 35-54 age group showed a non-significant reduction (aIRR=0.81, 95% CI: 0.62-1.04). Participants in households of 2–5 people had a 42% higher rate of infections than those living alone (aIRR=1.42, 95% CI:1.11–1.83).

### Sensitivity Analysis

When we examined work environment at different timepoints during the study, we found no difference in the model when replacing the three different work environment groups with the same variable measured during the baseline survey. (**Supplementary, Table S1**).

## DISCUSSION

In our study, age and household size emerged as the primary factors independently associated with COVID-19 infection rates. Younger adults (aged 18–34 years) and those living in medium-sized (2-5 person) households experienced significantly higher infection rates than people in single households after controlling for other sociodemographic and occupational factors.

The age association with COVID-19 infection in our study likely reflects differences in occupational and social exposures, with younger adults less able to work remotely and maintaining larger social networks. A large, pre-COVID-19 pandemic, multi-country modelling study explored how people of different ages mix in multiple settings (home, school, work), finding that social contacts are strongly age-assortative, where people mostly interact with their own age group, which has important implications for disease transmission patterns and control strategies.(14) Other studies have shown that during the COVID-19 pandemic younger adults were also more likely to engage in activities that increased virus transmission risk, such as dining indoors or attending social gatherings, and had a higher rate of non-adherence with isolation measures.(15, 16) Younger adults also faced greater occupational exposure to COVID-19 because they were employed in industries extensively disrupted by COVID-19 that could not easily pivot to remote work,(17) and were overrepresented in frontline occupations such as retail, hospitality, and entertainment resulting in sustained workplace exposure to infection.(18) In Australia, younger adults (aged 15-24 years) were less likely to work from home, as remote work was concentrated in high-skilled, white-collar jobs.(19)

In our study participants in households with two to five people had 42% higher infection rates compared to people in single-person households. We did not observe significantly increased infection rates in households with six or more people, likely due to the small sample size in that category (n=26). These findings align with other studies that have demonstrated an increasing risk of COVID-19 infection with increasing household size,(2, 20, 21) and underscores the critical role of household transmission in driving infection patterns, particularly during the first Omicron wave when most infections and deaths occurred in Australia.(6) Contrary to previous population-level studies that documented higher COVID-19 burdens among CALD and low-SES communities in Australia, neither CALD nor SES status independently predicted infection risk after adjustment for household size and age in our study, which may reflect a difference in study population and recruitment.(1, 2, 22, 23) Our study was not representative of the underlying population, rather aimed to recruit a high proportion of CALD participants; among the CALD group in our cohort 46% were high-SES compared to 54% low-SES.

Data from a 2022 report of housing conditions in Australia highlighted that low-income families and individuals from CALD communities, especially recently arrived migrants, are disproportionately affected by larger households and overcrowded living arrangements, in part because of housing affordability, inter-generational living patterns and cultural norms and obligations.(24) In the context of infectious disease transmission, such higher household density increases the risk of within-household spread and may partly explain the disproportionate COVID-19 burden observed in these communities, while also explaining why CALD status was not independently predictive in our cohort where CALD and non-CALD participants had similar household characteristics.

Our findings demonstrate that COVID-19 transmission patterns in our cohort were shaped primarily by age and household size rather than by cultural background or socioeconomic status as independent risk factors. The lack of independent CALD association after adjustment may reflect successful multilingual public health messaging in Victoria and equitable access to vaccines, though unmeasured confounders such as specific occupation types, job security and more granular data for socio-economic status warrant further investigation. Future pandemic preparedness should address the vulnerabilities of younger workers in essential and casual industries through workplace protections, paid sick leave policies and targeted isolation support. For larger households, public health responses could provide facilities for individuals with COVID-19 to isolate away from household members, financial support and incentives for effective quarantine and practical guidance on supporting multi-generational households where the index case may have dependants relying on their care. Reducing infection burden in future respiratory disease outbreaks requires policies that enable both young workers and those in higher density housing to protect themselves and their households without sacrificing economic security.

This study has several limitations. First, the cohort was predominantly female (75%), and as women generally demonstrated more protective behaviours during the pandemic,(25–27) this may have obscured associations between certain factors (such as CALD status, SES or healthcare worker status) and COVID-19 infection. Second, findings are limited by potential recall bias in self-reported infection history and by the definition of infection used, which relied on participant recall of positive tests or clinical diagnosis. Third, some baseline variables that impact COVID-19 infection risk, such as work from home status may have changed during the study period, though we explored this through the sensitivity analysis on working from home. Finally, the ‘non-CALD’ cohort in this study comprised a coalition of priority populations and therefore did not fully represent the overall non-CALD population in Victoria.

## CONCLUSION

This study identifies medium household size and younger age as the primary drivers of COVID-19 infection risk in this Victorian cohort, and contrary to previous literature, CALD and low-SES status did not independently predict infection risk after controlling for household size and age. These findings suggest that effective pandemic response requires moving beyond demographic categorisations of risk and toward addressing the material conditions that increase the risk of infection for individuals, such as housing size, workplace protections for casual workers, and economic security. As a relatively wealthy country, Australia has the opportunity in the current inter-pandemic period to address these structural factors and prevent repeating the inequitable infection outcomes we observed during the COVID-19 pandemic.

## Data Availability

All relevant data are within the manuscript and its Supporting Information files.

## Acknowledgements

Optimise is a partnership between Burnet Institute and Peter Doherty Institute in collaboration with University of Melbourne, Swinburne University of Technology, Monash University, La Trobe University, Murdoch Children’s Research Institute, the Centre for Culture Ethnicity and Health, and the Health Issues Centre. The authors gratefully acknowledge the generosity of the community members who participated in the study. The authors appreciatively acknowledge the work of all Optimise project team members and collaborators who have contributed to the ongoing delivery of the study. The authors gratefully acknowledge the contribution to this work of the Victorian Operational Infrastructure Support Program received by the Burnet Institute.

## REFERENCES

1. Roder C, Maggs C, McNamara BJ, O’Brien D, Wade AJ, Bennett C, et al. Area-level social and economic factors and the local incidence of SARS-CoV-2 infections in Victoria during 2020. Med J Aust. 2022;216(7):349–56.

2. Uribe Guajardo MG, Moore C, Giannopoulos V, Liu H, Tickle A, Adily P, et al. The impact of contextual socioeconomic and demographic characteristics of residents on COVID-19 outcomes during public health restrictions in Sydney, Australia. Aust N Z J Public Health. 2025;49(2):100228.

3. Australian Bureau of Statistics. COVID-19 Mortality in Australia, Deaths registered to 31 January 2022: ABS; 2022 [Available from: https://www.abs.gov.au/articles/covid-19-mortality-australia-deaths-registered-31-january-2022.

4. Australian Bureau of Statistics. COVID-19 Mortality in Australia: Deaths registered until 31 January 2024 2024 [Available from: https://www.abs.gov.au/articles/covid-19-mortality-australia-deaths-registered-until-31-january-2024.

5. Davey M, Nicholas, J. Covid death rate three times higher among migrants than those born in Australia. The Guardian. 2022 February 16, 2022.

6. Australian Bureau of Statistics. COVID-19 Mortality by wave: Australian Bureau of Statistics; 2022 [Available from: https://www.abs.gov.au/articles/covid-19-mortality-wave.

7. Pedrana A, Bowring A, Heath K, Thomas AJ, Wilkinson A, Fletcher-Lartey S, et al. Priority populations’ experiences of isolation, quarantine and distancing for COVID-19: protocol for a longitudinal cohort study (Optimise Study). BMJ Open. 2024;14(1):e076907.

8. Narayanasamy S, Altermatt A, Wilkinson AL, Heath K, Gibney K, Hellard M, et al. Adherence to Public Health Recommendations, Restrictions, and Requirements among Priority Populations at Risk for COVID-19 Mortality and Infection in Australia. medRxiv. 2026:2026.02.15.26346356.

9. Premier of Victoria. Changes To Pandemic Management: Victorian Government; 2022 [Available from: https://www.premier.vic.gov.au/changes-pandemic-management?utm_source=chatgpt.com.

10. Australian Broadcasting Cooperation. What do new changes to Victoria’s COVID-19 testing system mean for you?: Australian Broadcasting Cooperation; 2022 [Available from: https://www.abc.net.au/news/2022-01-06/changes-to-victoria-covid-testing-rules-rat-pcr/100741694?utm_source=chatgpt.com.

11. More Than Two Million Rapid Antigen Tests On The Way [press release]. Victorian Government 2021.

12. Australian Bureau of Statistics. Households more likely to test with RATs in March 2021 [Available from: https://www.abs.gov.au/media-centre/media-releases/households-more-likely-test-rats-march?utm_source=chatgpt.com.

13. Australian Bureau of Statistics. Socio-Economic Indexes for Areas (SEIFA) Australia 2021 [Available from: https://www.abs.gov.au/statistics/people/people-and-communities/socio-economic-indexes-areas-seifa-australia/latest-release.

14. Prem K, Cook AR, Jit M. Projecting social contact matrices in 152 countries using contact surveys and demographic data. PLoS Comput Biol. 2017;13(9):e1005697.

15. Moore RC, Lee AY, Hancock JT, Halley MC, Linos E. Age-Related Differences in Experiences With Social Distancing at the Onset of the COVID-19 Pandemic: A Computational and Content Analytic Investigation of Natural Language From a Social Media Survey. JMIR Hum Factors. 2021;8(2):e26043.

16. Murphy K, Williamson H, Sargeant E, McCarthy M. Why people comply with COVID-19 social distancing restrictions: Self-interest or duty? Australian & New Zealand Journal of Criminology. 2020;53(4):477–96.

17. Kabátek J. Jobless and distressed: the disproportionate effects of COVID-19 on young Australians. Australian Policy Online; 2020 23 September 2020.

18. Hâncean MG, Lerner J, Perc M, Ghiţă MC, Bunaciu DA, Stoica AA, et al. The role of age in the spreading of COVID-19 across a social network in Bucharest. J Complex Netw. 2021;9(4):cnab026.

19. Laβ I, Wooden M. Working From Home: The Australian Experience. Australian Economic Review. 2025;58(2):154–62.

20. Gillies CL, Rowlands AV, Razieh C, Nafilyan V, Chudasama Y, Islam N, et al. Association between household size and COVID-19: A UK Biobank observational study. J R Soc Med. 2022;115(4):138–44.

21. Chadeau-Hyam M, Wang H, Eales O, Haw D, Bodinier B, Whitaker M, et al. SARS-CoV-2 infection and vaccine effectiveness in England (REACT-1): a series of cross-sectional random community surveys. Lancet Respir Med. 2022;10(4):355–66.

22. Liu B, Spokes P, He W, Kaldor J. High risk groups for severe COVID-19 in a whole of population cohort in Australia. BMC Infect Dis. 2021;21(1):685.

23. Ward S, Restrepo AC, McHugh L. Area-level geographic and socioeconomic factors and the local incidence of SARS-CoV-2 infections in Queensland between 2020 and 2022. Aust N Z J Public Health. 2023;47(6):100094.

24. Dockery M, Moskos, Megan, Isherwood, Linda, Harris, Mark How many in a crowd? Assessing overcrowding measures in Australian housing. Australian Housing and Urban Research Institute; 2022.

25. Moran KR, Del Valle SY. A Meta-Analysis of the Association between Gender and Protective Behaviors in Response to Respiratory Epidemics and Pandemics. PLoS One. 2016;11(10):e0164541.

26. Galasso V, Pons V, Profeta P, Becher M, Brouard S, Foucault M. Gender differences in COVID-19 attitudes and behavior: Panel evidence from eight countries. Proc Natl Acad Sci U S A. 2020;117(44):27285–91.

27. Okten IO, Gollwitzer A, Oettingen G. Gender differences in preventing the spread of coronavirus. Behavioral Science & Policy. 2020;6(2):109–22.

